# Regional pleural strain measurements during mechanical ventilation using ultrasound elastography: A randomised, crossover, proof of concept physiologic study

**DOI:** 10.1101/2022.01.02.22268642

**Authors:** Martin Girard, Marie-Hélène Roy Cardinal, Michaël Chassé, Sébastien Garneau, Yiorgos Alexandros Cavayas, Guy Cloutier, André Y. Denault

**Affiliations:** Department of Anesthesiology, University of Montreal Hospital, Montréal, QC, Canada; Division of Critical Care, Department of Medicine, University of Montreal Hospital, Montréal, QC, Canada; University of Montreal Hospital Research Center, Montréal, QC, Canada; Laboratory of Biorheology and Medical Ultrasonics, University of Montreal Hospital Research Center, Montréal, QC, Canada; Department of Medicine, Sacré-Coeur Hospital of Montréal, Montréal, QC, Canada; Institute of Biomedical Engineering, University of Montreal, Montréal, QC, Canada; Department of Radiology, Radio-Oncology and Nuclear Medicine, University of Montreal, Montréal, QC, Canada; Department of Anesthesiology, Montreal Heart Institute, Montréal, QC, Canada

**Keywords:** General anesthesia, Lung imaging, Mechanical ventilation, Ultrasonography, Ventilator-Induced Lung Injury

## Abstract

**Background:** Mechanical ventilation is a common therapy in operating rooms and intensive care units. When ill-adapted, it can lead to ventilator-induced lung injury (VILI), which is associated with poor outcomes. Excessive regional pulmonary strain is thought to be a major mechanism responsible for VILI. Scarce bedside methods exist to measure regional pulmonary strain. We propose a novel way to measure regional pleural strain using ultrasound elastography.

**Research Question:** The objective of this study was to assess the feasibility and reliability of pleural strain measurement by ultrasound elastography and to determine if elastography parameters would correlate with varying tidal volumes.

**Study Design and Methods:** A single-blind randomized crossover proof of concept study was conducted July to October 2017 at a tertiary care referral center. Ten patients requiring general anesthesia for elective surgery were recruited. After induction, patients were received tidal volumes of 6, 8, 10 and 12 mL.kg^-1^ in random order, while pleural ultrasound cineloops were acquired at 4 standardized locations. Ultrasound radiofrequency speckle tracking allowed computing various pleural translation, strain and shear components. These were screened to identify those with the best dose-response with tidal volumes using linear mixed effect models. Goodness-of-fit was assessed by the coefficient of determination. Intraobserver, interobserver and test-retest reliability were calculated using intraclass correlation coefficients.

**Results:** Analysis was possible in 90.7% of ultrasound cineloops. Lateral *absolute* shear, lateral *absolute* strain and Von Mises strain varied significantly with tidal volume and offered the best dose-responses and data modelling fits. Point estimates for intraobserver reliability measures were excellent for all 3 parameters (0.94, 0.94 and 0.93, respectively). Point estimates for interobserver (0.84, 0.83 and 0.77, respectively) and test-retest (0.85, 0.82 and 0.76, respectively) reliability measures were good.

**Interpretation:** Strain imaging is feasible and reproducible, and may eventually guide mechanical ventilation strategies in larger cohorts of patients.

## Introduction

While often a live-saving therapy and a necessity during general anaesthesia, mechanical ventilation may promote lung damage when ill-adapted, a phenomenon called ventilator-induced lung injury (VILI)^1^. Some patient populations have been shown to be at higher risk of developing VILI: patients suffering from the acute respiratory distress syndrome^2^ and patients requiring one-lung ventilation^3^.

Assessing mechanical properties of the lung has been suggested as a promising approach to detect conditions leading to VILI^4^ with excessive pulmonary strain thought to be a critical factor in its development^5,6^. Unfortunately, pulmonary strain is non-trivial to measure at the bedside^7^ and most techniques measure global pulmonary strain, an oversimplification of the complex lung physiology. Areas of higher regional pulmonary strain that cannot be assessed using global measures have been implicated in the development of pulmonary inflammation^8^. Detecting these areas of higher regional pulmonary strain may enable identification of patients at higher risk of VILI who have seemingly benign global strain measures and yet may benefit from further strain-lowering interventions^9^.

While magnetic resonance imaging^10^, positron emission tomography^11^ and thoracic computed tomography^9^ are currently available in a research setting to measure regional pulmonary strain, these techniques are particularly burdensome, subject patients to the dangers of intra-hospital transport^12^, expose patients to ionizing radiations^13^ and haven’t been validated against a gold standard nor have they been shown to be associated with clinical outcome. Electrical impedance tomography is an increasingly popular monitoring modality in ventilated patients^14^ and has the advantage of being a bedside and radiation-free imaging modality. Unfortunately, electrical impedance tomography cannot currently measure regional pulmonary strain and requires costly dedicated equipment that is currently unavailable in most operating rooms and intensive care units. Consequently, all of the above modalities have major drawbacks that limit their use and validity.

Lung ultrasonography, a bedside, precise, repeatable, easy to learn and low-cost exam^15^, is already used to monitor pulmonary aeration during mechanical ventilation^16^. Preliminary works hint that it may be an interesting avenue to measure regional pleural strain as a surrogate for regional pulmonary strain^17,18^. Ultrasound (US) systems, now standard equipment in most operating rooms and intensive care, could make bedside regional pleural strain measurement commonplace. However, prospective feasibility data is lacking.

The objective of this study was to assess the feasibility and reliability (intraobserver, interobserver and test-retest) of pleural strain measurements by ultrasound elastography. Moreover, we sought to determine if elastography indices of pleural translation, strain and shear would correlate with varying tidal volumes.

## Methods

Detailed study procedures and analysis methods are described in the supplemental material. This report was redacted following the CONSORT 2010 extension to randomized crossover trials statement^19^.

### Study design

This study was a pilot single-centre, single-blind, randomized, four period crossover trial. A crossover design was chosen for this study to improve its power while dropouts and a potential carryover effect were not expected. The protocol was approved by the ethics committee of the Centre hospitalier de l’Université de Montréal (16.386) and registered at ClinicalTrials.gov (NCT03092557, registered March 28^th^, 2017). Written informed consent was obtained from all study participants.

### Study population

Between July and October 2017, adult patients with healthy lungs who were scheduled to undergo an elective surgery under general anaesthesia at the Centre hospitalier de l’Université de Montréal (Montreal, Canada), a tertiary care referral center, were screened for inclusion. To provide optimal imaging conditions for this pilot study, obese patients (body mass index > 30 kg.m^-2^) were excluded.

### Interventions

All patients were ventilated using volume-controlled ventilation, an inspired oxygen fraction of 40 to 50%, a respiratory rate of 12 min^-1^ and a positive end-expiratory pressure of 6 cm H_2_ O. After anesthesia induction, patients were administered tidal volumes of 6, 8, 10 and 12 mL.kg^-1^ predicted body weight ^20^ in random order (e-Figure 1). For each tidal volume, the pleura was imaged at 4 predetermined anatomical locations: left and right 3^rd^ intercostal space at the mid-clavicular line, and left and right 8^th^ intercostal space at the posterior axillary line. For each tidal volume and anatomical location tested, 3 ultrasound radiofrequency cineloops were acquired. Lung ultrasonography was performed using a Terason T3000cv scanner (Teratech Corporation, Burlington, MA) and a 12 MHz transducer (probe #12L5). The probe was oriented perpendicular to the ribs.

### Elastography

For each cineloop, the pleura was segmented manually on a single frame (e-Figure 2b). With the segmented pleura forming the upper boundary, a region of interest (ROI) of a fixed depth of 2 mm was defined (e-Figure 2c). The geometry of the ROI was automatically adapted and tracked throughout the respiratory cycle ^21^ or, if inadequate, simply copied over from frame to frame. Tracking was considered adequate when the pleural line remained within the ROI at all time. The Lagrangian speckle model estimator was used to compute tissue translation, strain and shear values (e-Figure 3) ^22^.

Elastography images and mechanical parameters were computed within the ROI over consecutive frames. Considering the planar nature of the pleura and its perpendicular orientation with respect to the US beam, we restricted our analysis to lateral translation, strain and shear components along with the Von Mises strain, a combination of bidimensional strain and shear components. Six elastography parameters were computed per cineloop (Figure 1, e-Table 1).

**Figure 1.**
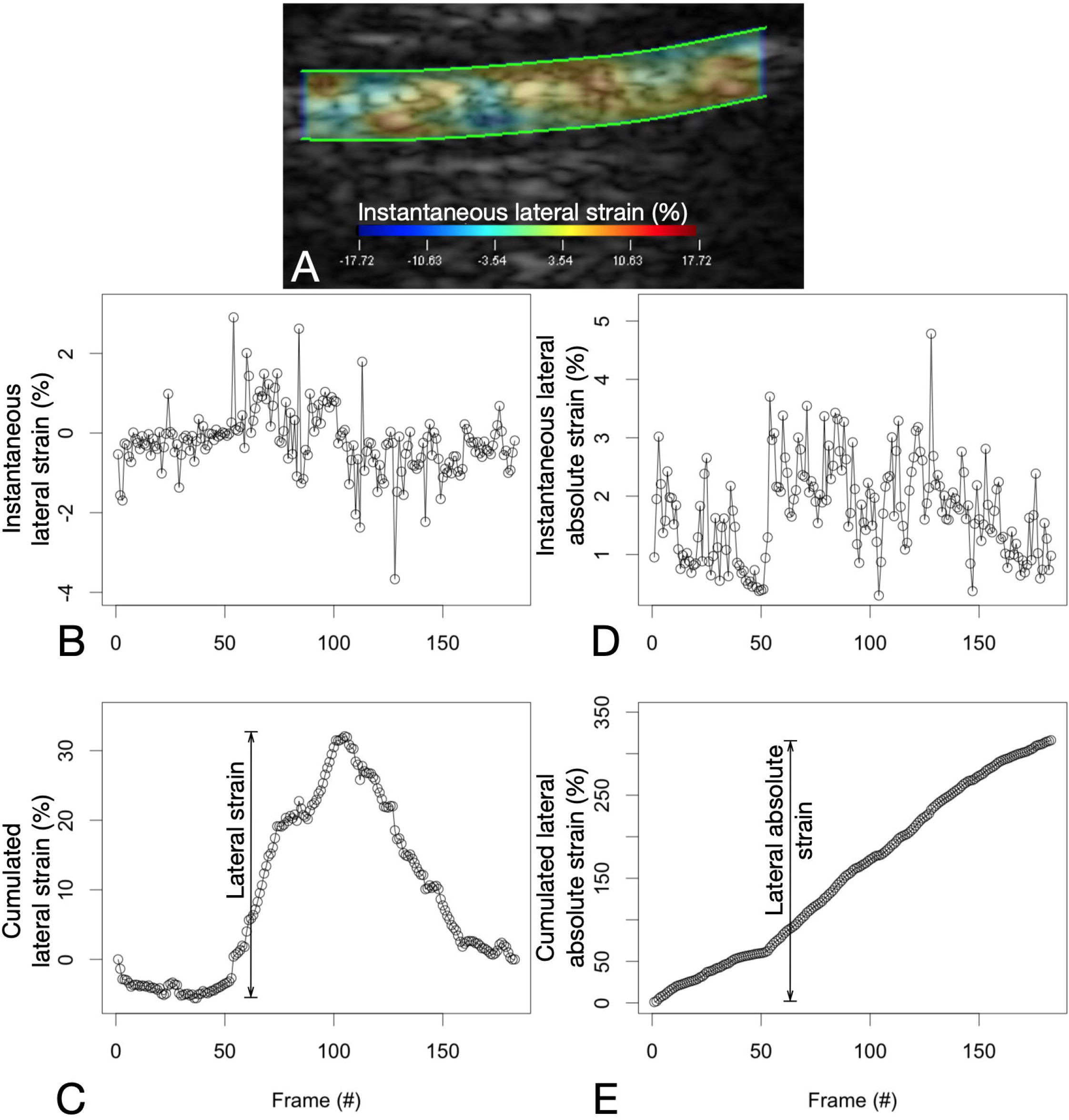
Calculating lateral strain and lateral *absolute* strain values. A: Instantaneous strain values are computed in all sub-ROIs between consecutive frames of a cineloop. B: By averaging all instantaneous sub-ROI strain values in a single frame, instantaneous strain values for the whole ROI are plotted for all frames of the cineloop. C: The summation of instantaneous strain values produces the cumulative strain of the pleura. Lateral strain is the range of the cumulative lateral strain experienced by the lung in the ROI. D: On the other hand, by averaging all *absolute* sub-ROI instantaneous strain values in a single frame, instantaneous *absolute* strain values for the whole ROI are plotted for all frames of the cineloop. E: The summation of the instantaneous *absolute* strain values produces the cumulative *absolute* strain of the pleura. Lateral *absolute* strain is the range of the cumulative lateral *absolute* strain experienced by the lung in the ROI.

### Randomization and blinding

Simple randomization was performed by a statistician not otherwise implicated in this study using a computer random number generator (R v3.4.0, R Core Team, 2017; blockrand package v1.3, Snow, 2013). Concealment was ensured by the use of sequentially numbered sealed and opaque envelopes.

### Objectives

The primary objective was the feasibility of pleural strain measurement using ultrasound elastography. Secondary objectives were: 1) verifying dose-response with varying tidal volumes and model fit for elastography parameters; 2) measuring reliability (intraobserver, interobserver and test-retest) of elastography parameters.

### Outcomes

The primary outcome was feasibility as defined by the number of cineloops on which elastography parameters were computed divided by the expected number of cineloops from the protocol.

### Statistical Analysis

We enrolled a convenience sample of 10 patients. For the primary outcome, with 72 potential cineloops per patient, our margin of error is 2.2% for a 95% confidence level and an expected 90% feasibility.

For the first secondary outcomes, linear mixed-effect models were used with elastography parameters as dependent variables and tidal volume, side of measurement and gravity dependence of measurement as independent variables. Interaction between tidal volume and gravity dependence was included in the model ^9^. All six slope estimates were tested for significance. A p value of 0.008 was considered significant. All other analyses are considered exploratory. Parameters with highest *absolute* values of estimated slopes with non-overlapping 95% confidence intervals were selected. Goodness of fit was assessed by the marginal and conditional coefficients of determination (R^2^)^23^. For the second secondary outcome, intraobserver, interobserver and test-retest reliability measures were calculated using intraclass correlation coefficients (ICC)^24^.

Results are expressed as mean ± standard deviation or median and interquartile range [25%-75%] as appropriate. Statistical analyses were performed using R (v3.4.0, R Core Team, 2017).

## Results

Twelve patients were assessed for eligibility. Two patients were excluded post-enrollment (e-Figure 4): one change in anesthetic plan and one consent withdrawal. Patient baseline characteristics are summarized in e-Table 2.

With our protocol specifying that 72 cineloops would be acquired for each of the 10 patients randomized, 720 elastograms were planned to be computed upon completion of cineloop acquisition. Sixty-two cineloops (8.6%) weren’t subjected to segmentation and tracking: 4 cineloops were not acquired because of a manipulation error, 7 cineloops were incomplete because of a technical issue with the echograph, and 51 cineloops didn’t show sufficient pleura throughout the respiratory cycle. This last problem was only encountered for cineloops acquired over the left 3^rd^ intercostal space at the mid-clavicular line where the heart was mostly seen. Lastly, elastograms were not computed for 6 cineloops because of inadequate tracking of the ROI throughout the respiratory cycle. Elastogram computation was thus performed on 652 (90.6%) cineloops.

Estimates for fixed effects of all 6 models can be found in Table 1. We observed a significant linear increase in 4 elastography parameters with increasing tidal volume: lateral *absolute* shear, lateral *absolute* strain, Von Mises strain and lateral *absolute* translation (Figure 2). With overlapping 95% confidence intervals, no significant elastography parameter was superior to the other in its ability to capture an increase in tidal volume (Figure 3). Amongst them, lateral *absolute* shear, lateral *absolute* strain and Von Mises strain were more precisely predicted by tidal volume compared to lateral *absolute* translation, as shown by higher goodness of fit (Table 1). The first sensitivity analysis yielded similar results (e-Table 3, e-Figure 5). When side and gravity dependence of measurements were considered, only gravity dependence seemed to be associated with our results. This suggests that pleural strain measurements in dependent lung zones may increase more rapidly with increasing tidal volume compared to nondependent zones. Amongst significant parameters, this seemed to be the case only for Von Mises strain (Figure 3, e-Figure 6).

**Table 1.** Modeled elastography parameters

| Elastography parameters | b<sub>1</sub> (slope) estimates for tidal volume |  | P value | Marginal R<sup>2</sup> | Conditional R<sup>2</sup> | Left effect estimates | Dependent effect estimates |
| --- | --- | --- | --- | --- | --- | --- | --- |
|  | Nondependent | Dependent | slope |  |  | (vs right) | (vs nondependent) |
| Lateral strain | 0.09<br>[-0.08 to 0.27] | 0.31<br>[0.15 to 0.47] | 0.3 | 0.20 | 0.48 | -0.43<br>[-0.78 to -0.08] | 0.64<br>[0.29 to 0.99] |
| Lateral translation | 0.09<br>[0.01 to 0.17] | 0.37<br>[0.3 to 0.45] | 0.03 | 0.46 | 0.89 | -0.24<br>[-0.63 to 0.15] | 1.25<br>[0.86 to 1.64] |
| Lateral absolute shear | 0.24<br>[0.16 to 0.33] | 0.37<br>[0.3 to 0.45] | < 0.0001 | 0.39 | 0.89 | 0.04<br>[-0.37 to 0.44] | -1.13<br>[-1.54 to -0.73] |
| Lateral absolute strain | 0.25<br>[0.17 to 0.34] | 0.41<br>[0.34 to 0.49] | < 0.0001 | 0.37 | 0.88 | 0.07<br>[-0.36 to 0.49] | -1.05<br>[-1.48 to -0.63] |
| Von Mises strain | 0.27 | 0.51 | < 0.0001 | 0.40 | 0.86 | 0.07 | -1.01 |
|  | [0.18 to 0.37] | [0.42 to 0.6] |  |  |  | [-0.35 to 0.49] | [-1.44 to -0.59] |
| Lateral absolute translation | 0.3<br>[0.17 to 0.43] | 0.55<br>[0.42 to 0.67] | < 0.0001 | 0.21 | 0.72 | 0.26<br>[-0.2 to 0.72] | -0.23<br>[-0.69 to 0.23] |

Ordered by increasing b1 (slope) estimate.

**Figure 2.**
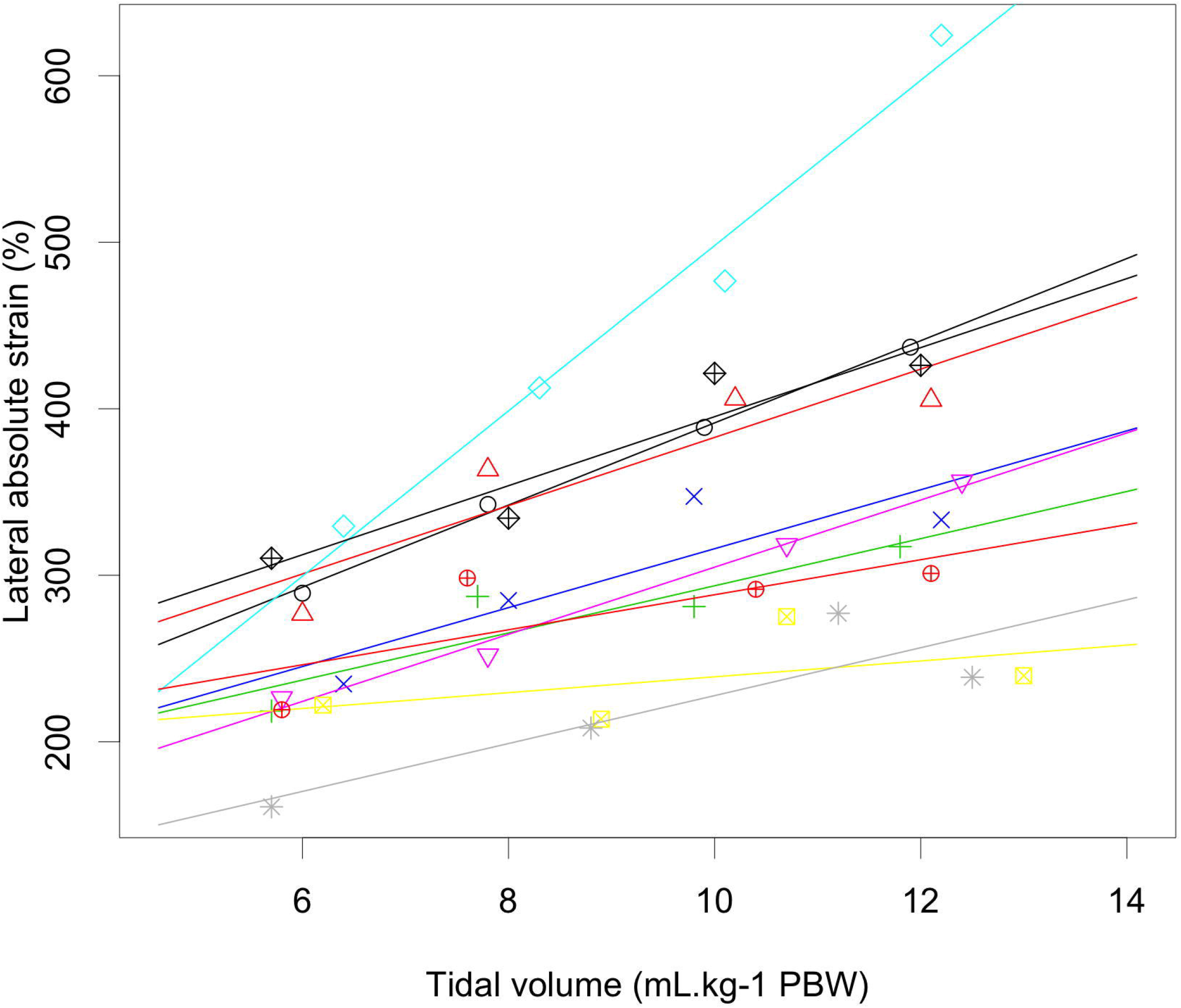
Regression lines for the lateral *absolute* strain across various tidal volumes administered with each color and symbol representing a different patient. Cineloops were acquired over the left 8^th^ intercoastal space at the posterior axillary line. PBW: predicted body weight.

**Figure 3.**
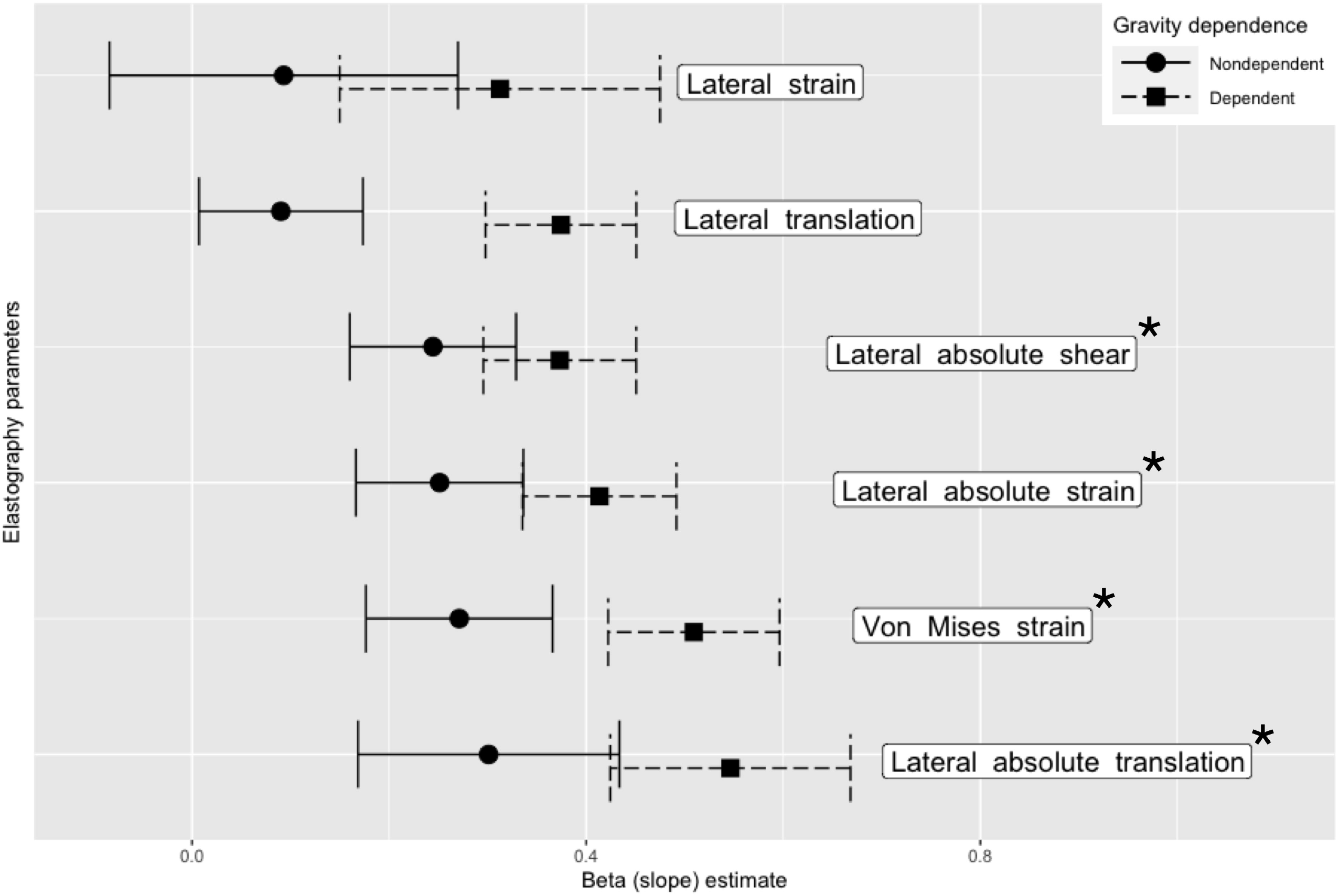
Slope estimates for elastography parameters in increasing order stratified by gravity dependence. Significant parameters are identified by an Asterix.

Measured intraobserver reliability was excellent for lateral *absolute* shear, lateral *absolute* strain and Von Mises strain. Interobserver and test-retest measured reliabilities were found to be moderate to good from Von Mises strain and moderate to excellent for lateral *absolute* shear and lateral *absolute* strain (Figure 4, e-Table 4). In our sensitivity analysis, bootstrapping yielded identical estimates (e-Table 5).

**Figure 4.**
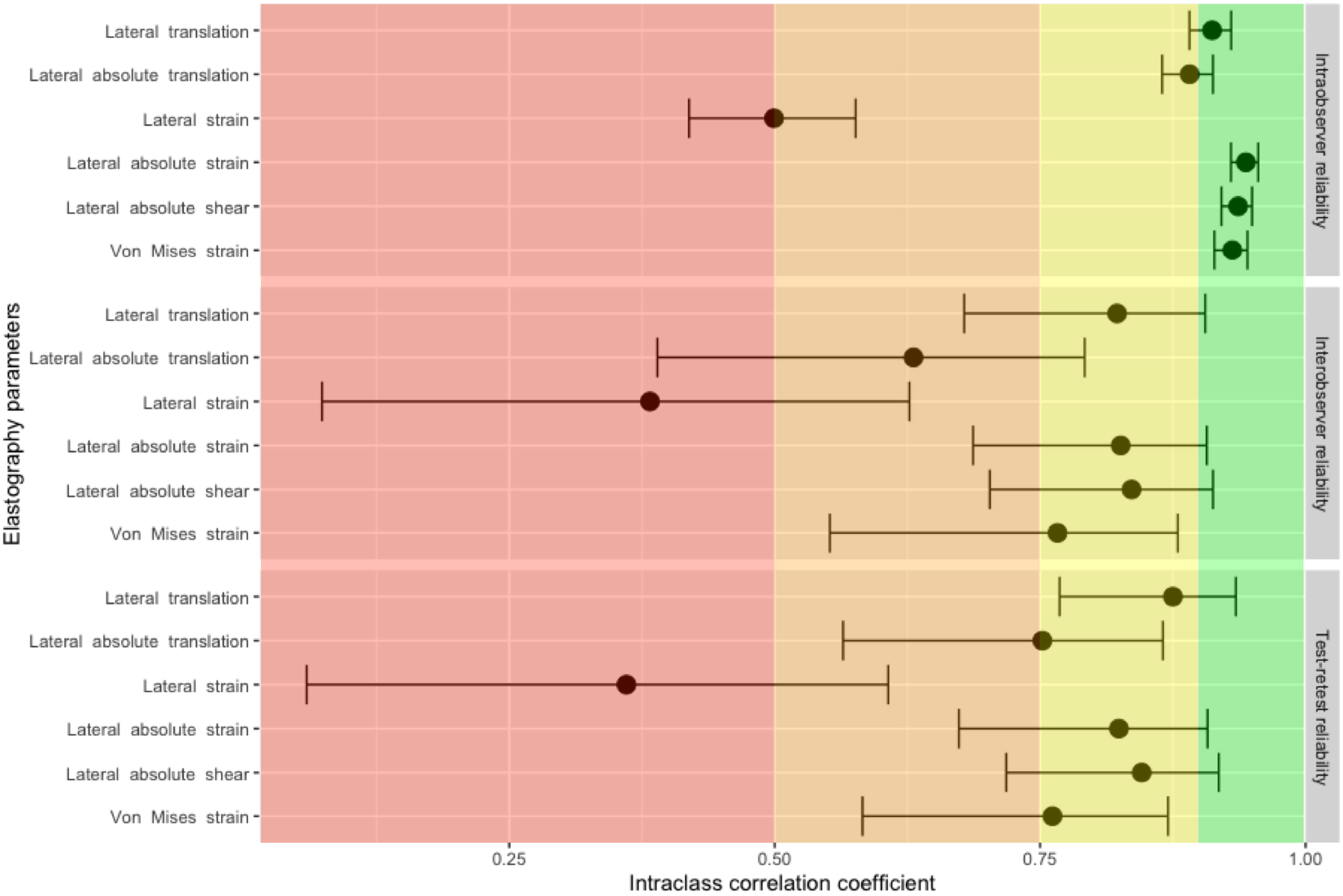
Intraclass correlation coefficients and 95% confidence intervals for intraobserver, interobserver and test-retest reliability measures for elastography parameters. Intraclass correlation coefficients in the red panel indicate poor reliability, values in the orange panel indicate moderate reliability, values in the yellow panel indicate good reliability and values in the green panel indicate excellent reliability.

## Discussion

We showed preliminary evidence that computing regional pleural translation, strain and shear components is feasible in over 90% of cineloops. Lateral *absolute* shear, lateral *absolute* strain and Von Mises strain were more informative in regards to administered tidal volumes than lateral translation, lateral *absolute* translation and lateral strain. Measured intraobserver reliability was excellent for all significant parameters while interobserver and test-retest measured reliability were found to be moderate to excellent. This supports our hypothesis that lung ultrasound could be used to measure regional strain in mechanically ventilated patients.

While others have previously described ultrasonographically-measured regional pleural strain in humans ^17,18^, this study significantly extends previous work by quantifying the impact of tidal volume on regional pleural strain. Despite the absence of a gold standard precluding any firm conclusion about the utility of each elastography parameter, we described an expected dose-response relationship between various strain measurements and administered tidal volumes.

At first glance, it is somewhat surprising that a 2 mm thick ROI delineated from an image generated by a 12 MHz probe could capture physiological signal from the pleura, a structure only 30 to 40 microns thick^25^. With the topmost part of the ROI aligned with the pleura, the image contained within the ROI is actually mainly composed of reverberation artefacts originating from the pleura and other soft tissue/ultrasound beam interfaces^26^. These artefacts are most easily seen when contrasting normal lung sliding in M-mode and their disappearance when a pneumothorax occurs (e-Figure 7). We hypothesize that reverberation artifacts in the ROI convey anatomical information on the pleura thus allowing our algorithm to compute elastography parameters. Another possibility would be that imperfect ROI tracking leads to intercoastal muscle being included in the strain calculation. While it is technically possible to measure intercostal muscle strain^27^, we do not believe this is the case with our results as patients were paralysed for induction of general anesthesia.

One important finding is that lateral strain had a poor dose-response with tidal volume despite a time plot of lateral strain that seemed to track delivered tidal volume in many patients (Figure 1C). Perhaps explaining the poor performance of lateral strain compared to lateral *absolute* strain, we observed small areas of negative strain during lung inflation (Figure 1A). Supporting this hypothesis, zones of local deflation during inspiration have been described in animals (e-Figure 8)^28,29^. Analogous to how lateral strain and lateral *absolute* strain are calculated, global pulmonary compliance was correlated with the sum of positive values of local compliance, measured by computed tomography, but not with the sum of positive and negative values of local compliance^29^. While the exact causes for this phenomenon are still debated, tidal recruitment with gas redistribution may contribute. Discrepancies between lateral strain and lateral *absolute* strain could signal tidal recruitment and ongoing atelectrauma.

Our study has some important strengths and limitations. First, our results are physiologically plausible with lateral *absolute* shear, lateral *absolute* strain and Von Mises strain increasing with increasing tidal volumes. Second, our two sensitivity analyses demonstrate the robustness of results. Third, by performing a randomized crossover study, we ruled out the possibility of an order effect. Fourth, we also demonstrated the good to excellent reliability of these measurements. Given the proof-of-concept nature of this study, several limitations should be considered. First, the limited sample size restricts our ability to obtain precise estimates of pleural strain in human. Second, no gold standard for pulmonary strain was available for comparison. While helium dilution or nitrogen-washout techniques could have been used to measure end-expiratory lung volume at each step of our protocol, these methods cannot account for tidal recruitment or overdistension^30^. Third, whether regional pleural strain is a good surrogate for regional pulmonary strain remains to be determined. Fourth, flow, pressure and volume curves were not recorded simultaneously with each image acquisition. As such, the precise beginning and end of each respiratory cycle wasn’t exactly known which may have affected our results (Figure 2e). The exact tidal volume administered while acquiring cineloops may also have been slightly different from the tidal volume recorded beforehand. Finally, physiological lung sliding and rib shadows preclude measuring the same pleural surface throughout the respiratory cycle. While we computed strain between two consecutive frames of a given cineloop, the pleura imaged in the ROI at end-inspiration was likely different from the pleura in the ROI at end-expiration. As such, strain measures likely represent average strain values from a pulmonary neighbourhood.

In conclusion, measuring regional pleural strain by ultrasound elastography is feasible. A significant dose-response relationship between tidal volume and lateral *absolute* shear, lateral *absolute* strain and Von Mises strain was observed, supporting the hypothesis that regional pulmonary strain can be measured at the bedside with non-invasive lung ultrasound. Further work will be required to compare elastography parameters to gold-standard strain measurements and how they can be used to tailor mechanical ventilation in the operating room and the intensive care unit to improve patient outcomes.

## Supporting information

CONSORT checklist

## Data Availability

All data produced in the present study are available upon reasonable request to the authors.

## Abbreviation List

ICC: intraclass correlation coefficients
R^2^: coefficient of determination
ROI: region of interest
US: ultrasound
VILI: ventilator-induced lung injury

## Guarantor statement

MG had full access to all of the data in the study and takes responsibility for the integrity of the data and the accuracy of the data analysis, including and especially any adverse effects.

## Author contributions

M.G.: Study design, data collection, data analysis, writing first draft of paper and revising final draft of paper.

M.H.R.C.: Data analysis and revising final draft of paper.

M.C.: Data analysis and revising final draft of paper.

S.G.: Data collection and revising final draft of paper.

Y.A.C.: Study design and revising final draft of paper.

G.C.: Study design, data analysis and revising final draft of paper.

A.D.: Study design, data analysis and revising final draft of paper.

## Data availability statement

All data produced in the present study are available upon reasonable request to the corresponding author.

## Other contributions

The authors thank Ms. Monique Ruel, RN, for her valuable assistance and dedication and Dr Brian Grondin-Beaudoin, MD, for his help with preliminary testing procedures.

