## Supplementary material for "Regional pleural strain measurements during mechanical ventilation using ultrasound elastography: A randomised, crossover, proof of concept physiologic study": CONSORT checklist

| CONSORT checklist of information to include when reporting randomised crossover trials | | | |
| --- | --- | --- | --- |
| Section/topic | Item No | Description | Page No* |
| Title† | 1a | Identification as a randomised crossover trial in the title | 1 |
| Abstract† | 1b | Specify a crossover design and report all information outlined in table 2 | 3-4 |
| Introduction: | | | |
| Background‡ | 2a | Scientific background and explanation of rationale | 6-7 |
| Objectives‡ | 2b | Specific objectives or hypotheses | 7 |
| Methods: | | | |
| Trial design† | 3a | Rationale for a crossover design. Description of the design features including allocation ratio, especially the number and duration of periods, duration of washout period, and consideration of carry over effect | 8 |
| Change from protocol‡ | 3b | Important changes to methods after trial commencement (such as eligibility criteria), with reasons | NA |
| Participants‡ | 4a | Eligibility criteria for participants | 8 & SM |
| Settings and location‡ | 4b | Settings and locations where the data were collected | 8 |
| Interventions† | 5 | The interventions with sufficient details to allow replication, including how and when they were actually administered | 8-9 & SM |
| Outcomes‡ | 6a | Completely defined prespecified primary and secondary outcome measures, including how and when they were assessed | 11 & SM |
| Changes to outcomes‡ | 6b | Any changes to trial outcomes after the trial commenced, with reasons | NA |
| Sample size† | 7a | How sample size was determined, accounting for within participant variability | 11 & SM |
| Interim analyses and stopping guidelines‡ | 7b | When applicable, explanation of any interim analyses and stopping guidelines | NA |
| Randomisation: | | | |
| Sequence generation‡ | 8a | Method used to generate the random allocation sequence | 10 |
| Sequence generation‡ | 8b | Type of randomisation; details of any restriction (such as blocking and block size) | 10 |
| Allocation concealment mechanism‡ | 9 | Mechanism used to implement the random allocation sequence§ (such as sequentially numbered containers), describing any steps taken to conceal the sequence until interventions were assigned | 10 |
| Implementation† | 10 | Who generated the random allocation sequence,§ who enrolled participants, and who assigned participants to the sequence of interventions | 8-10 |
| Blinding‡ | 11a | If done, who was blinded after assignment to interventions (for example, participants, care providers, those assessing outcomes) and how | 10 |
| Similarity of interventions‡ | 11b | If relevant, description of the similarity of interventions | NA |
| Statistical methods† | 12a | Statistical methods used to compare groups for primary and secondary outcomes which are appropriate for crossover design (that is, based on within participant comparison) | 11-12 & SM |
| Additional analyses‡ | 12b | Methods for additional analyses, such as subgroup analyses and adjusted analyses | 11-12 & SM |
| Results | | | |
| Participant flow (a diagram is strongly recommended)† | 13a | The numbers of participants who were randomly assigned, received intended treatment, and were analysed for the primary outcome, separately for each sequence and period | SM |
| Losses and exclusions† | 13b | No of participants excluded at each stage, with reasons, separately for each sequence and period | 13 & SM |
| Recruitment‡ | 14a | Dates defining the periods of recruitment and follow-up | 8 |
| Trial end‡ | 14b | Why the trial ended or was stopped | 8 |
| Baseline data† | 15 | A table showing baseline demographic and clinical characteristics by sequence and period | SM |
| Numbers analysed† | 16 | Number of participants (denominator) included in each analysis and whether the analysis was by original assigned groups | 13 |
| Outcomes and estimation† | 17a | For each primary and secondary outcome, results including estimated effect size and its precision (such as 95% confidence interval) should be based on within participant comparisons.¶ In addition, results for each intervention in each period are recommended | 13-14 & SM |
| Binary outcomes‡ | 17b | For binary outcomes, presentation of both absolute and relative effect sizes is recommended | NA |
| Ancillary analyses‡ | 18 | Results of any other analyses performed, including subgroup analyses and adjusted analyses, distinguishing prespecified from exploratory | 14 & SM |
| Harms† | 19 | Describe all important harms or untended effects in a way that accounts for the design (for specific guidance, see CONSORT for harms^32^) | NA |
| Discussion: | | | |
| Limitations† | 20 | Trial limitations, addressing sources of potential bias, imprecision, and if relevant, multiplicity of analyses. Consider potential carry over effects | 17-18 |
| Generalisability‡ | 21 | Generalisability (external validity, applicability) of the trial findings | 18 |
| Interpretation‡ | 22 | Interpretation consistent with results, balancing benefits and harms, and considering other relevant evidence |  |
| Other information: | | | |
| Registration‡ | 23 | Registration number and name of trial registry |  |
| Protocol‡ | 24 | Where the full trial protocol can be accessed, if available | NA |
| Funding‡ | 25 | Sources of funding and other support (such as supply of drugs), role of funders |  |
