## Supplemental material for "Regional pleural strain measurements during mechanical ventilation using ultrasound elastography: A randomised, crossover, proof of concept physiologic study"

**Supplementary Materials & Methods**

Study population

Between July and October 2017, adult patients with healthy lungs who were scheduled to undergo an elective surgery under general anaesthesia requiring endotracheal intubation and muscle relaxation were screened for inclusion. Patients were recruited at the Centre hospitalier de l’Université de Montréal (Montreal, Canada), a tertiary care referral center. Patients with healthy lungs were defined as: no active or past history of smoking, no previous intrathoracic procedure, no known pulmonary disease, no oxygen requirement and metabolic equivalent task (METS) greater than or equal to 4. To provide optimal imaging conditions for this pilot study, obese patients (body mass index > 30 kg.m^-2^) were excluded.

Interventions

All patients were pre-oxygenated in the supine position with 100% oxygen for 3 minutes without any continuous positive airway pressure. General anesthesia induction was performed using standard doses of propofol and fentanyl. Rocuronium was used in all cases to facilitate tracheal intubation. Anesthesia was maintained with desflurane or sevoflurane. All patients were ventilated with Datex-Ohmeda Aestiva 3000 machines (GE Healthcare, WI, USA) using volume-controlled ventilation, an inspired oxygen fraction (F_i_O_2_) of 40 to 50%, a respiratory rate of 12 min^-1^, an inspiratory to expiratory ratio of 1:2, no inspiratory pause and a positive end-expiratory pressure of 6 cm H_2_O.

After anesthesia induction, patients were administered tidal volumes of 6, 8, 10 and 12 mL.kg^-1^ predicted body weight^1^ in random order (Figure E1). For each tidal volume, mean expired tidal volume of 3 consecutive breaths were collected and the pleura was imaged at 4 predetermined anatomical locations: left and right 3^rd^ intercostal space at the mid-clavicular line, and left and right 8^th^ intercostal space at the posterior axillary line. The correct intercostal spaces were identified by sliding the ultrasound transducer from the clavicle downwards and visually counting rib spaces. For each tidal volume and anatomical location tested, 3 ultrasound radiofrequency cineloops at a frame rate of 30 Hz were acquired over 3 separate respiratory cycles. All cineloops were saved to digital format for offline analysis. Interobserver reliability was assessed by repeating cineloop acquisition by a second blinded observer (SG) for the 10 mL.kg^-1^ predicted body weight tidal volume. Immediately after, test-retest reliability was also assessed by repeating cineloop acquisition by the first observer (MG) for the 10 mL.kg^-1^ predicted body weight tidal volume.

Lung ultrasonography

Lung ultrasonography was performed by experienced lung echographists (MG and SG for repeated measures with 8 and 1 years of experience) using a Terason T3000cv scanner (Teratech Corporation, Burlington, MA) and a 12 MHz transducer (probe #12L5). Initial depth of field was 4 cm and adjusted as needed to position the pleura between half to three-quarters of the screen. A single focal zone was placed nearest to the pleura. With the marker pointing towards the head, the probe was oriented perpendicular to the ribs with the pleura as horizontal as possible.

Elastography

B-mode images were reconstructed from radiofrequency data (Figure E2a). For each cineloop, the pleura was segmented manually on a single frame (Figure E2b). With the segmented pleura forming the upper boundary, a region of interest (ROI) of a fixed depth of 2 mm was defined (Figure E2c). The geometry of the ROI was automatically adapted and tracked throughout the respiratory cycle^2^ or, if inadequate, simply copied over from frame to frame. Tracking was considered adequate when the pleural line remained within the ROI at all time as assessed visually by an experienced lung echographist (MG). The Lagrangian speckle model estimator was used to compute tissue translation, strain and shear values (Figure E3)^3^. We defined translations as rigid displacements produced by lung sliding, strain as expansion or contraction of the pleura from tidal volume administration and shear as the angular deformation.

Elastography images and mechanical parameters were computed within the ROI over consecutive frames. We used an implementation of the estimator integrated into a commercial imaging platform (Visual, Object Research Systems)^4^. Axial and lateral elastography components were determined (“axial” indicating the direction along the US beam and “lateral” indicating the direction perpendicular to it). Considering the planar nature of the pleura and its perpendicular orientation with respect to the US beam, we restricted our analysis to lateral translation, strain and shear components along with the Von Mises strain, a combination of bidimensional strain and shear components. Six elastography parameters were computed per cineloop of a given patient (Figure 1, Table E1).

Outcomes

The primary outcome was feasibility as defined by the number of cineloops on which elastography parameters were computed divided by the expected number of cineloops from the protocol. Secondary outcomes were: 1) estimated elastography parameter slopes for tidal volume and marginal and conditional coefficients of determination (R^2^); 2) intraclass correlation coefficients (ICC) for intraobserver, interobserver and test-retest reliability values of elastography parameters.

Statistical Analysis

We enrolled a convenience sample of 10 patients. For the primary outcome, with 72 potential cineloops per patient, our margin of error is 2.2% for a 95% confidence level and an expected 90% feasibility.

For the first secondary outcomes, computation results of all 6 elastography parameters were modelled. All continuous dependent and independent variables were centered and reduced. This allowed direct comparison of slope estimates from the various models. To simplify models, all repeated measurements performed at a tidal volume of 10 mL.kg^-1^ used to measure interobserver and test-retest reliability values were not included in the models. Linear mixed-effect models were used with elastography parameters as dependent variables and tidal volume, side of measurement (left/right) and gravity dependence of measurement (dependent/nondependent) as independent variables. Because of the repeated (and thus correlated) nature of measurements per patient and per anatomical location, a random intercept per patient was included in the model as well as a one per anatomical location as a nested grouping factor, whereas triplicate measurements used to measure intraobserver reliability were averaged. Interaction between tidal volume and gravity dependence was included in the model ^5^. Model assumptions were verified. All six slope estimates were tested for significance. Using Bonferroni’s adjustment, a *p* value of 0.008 (0.05 / 6) was considered significant. All other analyses are considered exploratory. To identify elastography parameters with the best dose-response with tidal volumes, absolute values of estimated slopes for significant parameters were ordered. Parameters with highest absolute values of estimated slopes with non-overlapping 95% confidence intervals were selected. Goodness of fit was assessed by the marginal and conditional R^2^ ^6^. To ensure robustness of results, a sensitivity analysis was performed by using the same linear mixed-effect models described above but using a smaller subset of cineloops with better imaging quality. Cineloops with better imaging quality were defined as having a thin and clearly defined pleural line and perfect ROI tracking.

For the second secondary outcome, intraobserver, interobserver and test-retest reliability measures were calculated using ICC type (2,1)^7^. As outlined above, cineloops were acquired by different observers while the analysis process was performed by the same one. Bootstrap was performed as a second sensitivity analysis to calculate 95% confidence intervals using 10,000 iterations and the bias-corrected and accelerated method^8^. As suggested, ICC values less than 0.5 indicate poor reliability, values between 0.5 and 0.75 indicate moderate reliability, values between 0.75 and 0.9 indicate good reliability and values greater than 0.9 indicate excellent reliability^7^.

Results are expressed as mean ± standard deviation or median and interquartile range [25%-75%] as appropriate. No imputation for missing values was performed. Statistical analyses were performed using R (v3.4.0, R Core Team, 2017).

**Supplementary Figure legends**

**e- Figure 1. Example schematic diagram of study protocol and interventions in a sample patient.**

**e-Figure 2. Pleura segmentation process.** A: All cineloops are reconstructed from radiofrequency data. B: The pleura is segmented manually on a single frame. C: A region of interest of a fixed depth of 2 mm is defined with the segmented pleura forming the upper boundary.

**e-Figure 3. Schematic representation of tissue axial and lateral translation, strain and shear.**

**e-Figure 4. Study inclusion/exclusion flow diagram.**

**e-Figure 5. Slope estimates for elastography parameters** in increasing order stratified by gravity dependence for the first sensitivity analysis. Significant parameters are identified by an asterix.

**e-Figure 6. Regression lines and individual data points for Von Mises strai**n across the various tidal volumes stratified by gravity dependence. PBW: predicted body weight

**e-Figure 7. Reverberation artefacts (white asterix) generated by the visceral pleura** during normal lung sliding lead to the seashore sign when imaged in M-mode. When a pneumothorax occur, loss of reverberation from the separation of the parietal and visceral pleura leads to the barcode sign. The transition between both states (white arrow) is called a lung point

**e-Figure 8. Example compliance map during lung inflation in a heathy pig under general anesthesia at PEEP of 5 cm H_2_O**. Areas of regional lung inflation are colored in red while areas of regional lung deflation are colored in turquoise. Modified after Perchiazzi G, Rylander C, Derosa S, et al. *Respir Physiol Neurobiol*. 2014;201:60-70

**e-Table 1. Description of elastography parameters**

| **Parameters (units)** | **Description** |
| --- | --- |
| **Lateral** |  |
| Shift |  |
| Lateral translation (mm) | Range of the cumulated lateral shift. It represents the range of the distance travelled by the pleura on both sides of its starting point because of lung sliding. |
| Lateral absolute translation (mm) | Range of the absolute cumulated lateral shift. The absolute cumulated lateral shift was calculated by summation of a per-frame average of the absolute values of all individual sub-ROI computed instantaneous lateral shifts. It is always positive and represents the total distance travelled by the pleura throughout the respiratory cycle because of lung sliding. |
| Strain |  |
| Lateral strain (%) | Range of the cumulated lateral strain. It represents the range of the expansion (or contraction) of the pleura from tidal volume insufflation and exsufflation. |
| Lateral absolute strain (%) | Range of the absolute cumulated lateral strain. The absolute cumulated lateral strain was calculated by summation of a per-frame average of the absolute values of all individual sub-ROI computed instantaneous lateral strain. It is always positive and represents the total lateral strain (expansion and contraction) experienced by the pleura throughout the respiratory cycle from tidal volume insufflation and exsufflation. |
| Shear |  |
| Lateral absolute shear (%) | Range of the absolute cumulated lateral shear. The absolute cumulated lateral shear was calculated by summation of a per-frame average of the absolute values of all individual sub-ROI computed instantaneous lateral shear. It is always positive and represents the total angular strain (left-sided and right-sided) experienced by the pleura throughout the respiratory cycle from tidal volume insufflation and exsufflation. |
| **Bidimensional** |  |
| Von Mises strain (%) | Range of the cumulated Von Mises strain. Von Mises strain is a combination of axial and lateral strain and shear components. It is always positive and represents the magnitude of the total strain experienced by the pleura throughout the respiratory cycle from tidal volume insufflation and exsufflation. |

**e-Table 2. Patient characteristics**

| Variables | Value (n = 10) |
| --- | --- |
| Age (y) | 53 [37 – 66] |
| Sex, M/F (no) | 5/5 |
| ASA classification, 1/2/3 (no) | 3/5/2 |
| Height (cm) | 167 [163 – 177] |
| Weight (kg) | 74 [64 – 89] |
| Body mass index (kg.m^-2^) | 27 [24 – 30] |
| Predicted Body Weight (kg) | 61 [55 – 72] |

All data presented as median [interquartile range] unless otherwise specified.

**e-Table 3. Modeled elastography parameters using reduced dataset**

| Elastography parameters | b_1_ (slope) estimates | | P value | Marginal  R^2^ | Conditional  R^2^ | Left effect estimates | Dependent effect estimates |
| --- | --- | --- | --- | --- | --- | --- | --- |
|  | Nondependent | Dependent | slope |  |  | (vs right) | (vs nondependent) |
| Lateral strain | 0.14  [-0.07 to 0.35] | 0.38  [0.21 to 0.55] | 0.2 | 0.20 | 0.48 | -0.33  [-0.72 to 0.05] | 0.63  [0.24 to 1.02] |
| Lateral translation | 0.11  [0.01 to 0.21] | 0.4  [0.32 to 0.48] | 0.03 | 0.42 | 0.89 | -0.17  [-0.63 to 0.3] | 1.2  [0.73 to 1.67] |
| Lateral absolute shear | 0.33  [0.23 to 0.42] | 0.39  [0.32 to 0.47] | < 0.0001 | 0.43 | 0.90 | 0.1  [-0.36 to 0.57] | -1.16  [-1.63 to -0.69] |
| Lateral absolute strain | 0.33  [0.24 to 0.42] | 0.43  [0.35 to 0.5] | < 0.0001 | 0.40 | 0.91 | 0.15  [-0.32 to 0.62] | -1.07  [-1.54 to -0.6] |
| Von Mises strain | 0.33  [0.23 to 0.43] | 0.51  [0.43 to 0.59] | < 0.0001 | 0.44 | 0.89 | 0.15  [-0.33 to 0.63] | -1.06  [-1.53 to -0.58] |
| Lateral absolute translation | 0.32  [0.18 to 0.46] | 0.52  [0.4 to 0.63] | < 0.0001 | 0.23 | 0.77 | 0.38  [-0.15 to 0.9] | -0.34  [-0.86 to 0.19] |

Parameters ordered as in Table 1 from the main manuscript for easier comparison.

**e-Table 4. Intraobserver, interobserver and test-retest measured reliability for elastography parameters**

| Elastography parameters | Intraobserver  reliability | Interobserver  reliability | Test-retest  reliability |
| --- | --- | --- | --- |
| Lateral translation | 0.91 [0.89 to 0.93] | 0.82 [0.68 to 0.91] | 0.88 [0.77 to 0.93] |
| Lateral absolute translation | 0.89 [0.86 to 0.91] | 0.63 [0.39 to 0.79] | 0.75 [0.56 to 0.87] |
| Lateral strain | 0.5 [0.42 to 0.58] | 0.38 [0.07 to 0.63] | 0.36 [0.06 to 0.61] |
| Lateral absolute strain | 0.94 [0.93 to 0.96] | 0.83 [0.69 to 0.91] | 0.82 [0.67 to 0.91] |
| Lateral absolute shear | 0.94 [0.92 to 0.95] | 0.84 [0.7 to 0.91] | 0.85 [0.72 to 0.92] |
| Von Mises strain | 0.93 [0.91 to 0.95] | 0.77 [0.55 to 0.88] | 0.76 [0.58 to 0.87] |

**e-Table 5. Intraobserver, interobserver and test-retest measured reliability with bootstrapped 95% confidence intervals for elastography parameters**

| Elastography parameters | Intraobserver  reliability | Interobserver  reliability | Test-retest  reliability |
| --- | --- | --- | --- |
| Lateral translation | 0.91 [0.88 to 0.93] | 0.82 [0.69 to 0.9] | 0.88 [0.76 to 0.93] |
| Lateral absolute translation | 0.89 [0.86 to 0.92] | 0.63 [0.38 to 0.8] | 0.75 [0.52 to 0.86] |
| Lateral strain | 0.5 [0.43 to 0.57] | 0.38 [0.17 to 0.56] | 0.36 [-0.08 to 0.63] |
| Lateral absolute strain | 0.94 [0.92 to 0.96] | 0.83 [0.65 to 0.91] | 0.82 [0.63 to 0.89] |
| Lateral absolute shear | 0.94 [0.92 to 0.95] | 0.84 [0.68 to 0.91] | 0.85 [0.72 to 0.9] |
| Von Mises strain | 0.93 [0.91 to 0.95] | 0.77 [0.56 to 0.88] | 0.76 [0.55 to 0.87] |

**e-Figure 1**

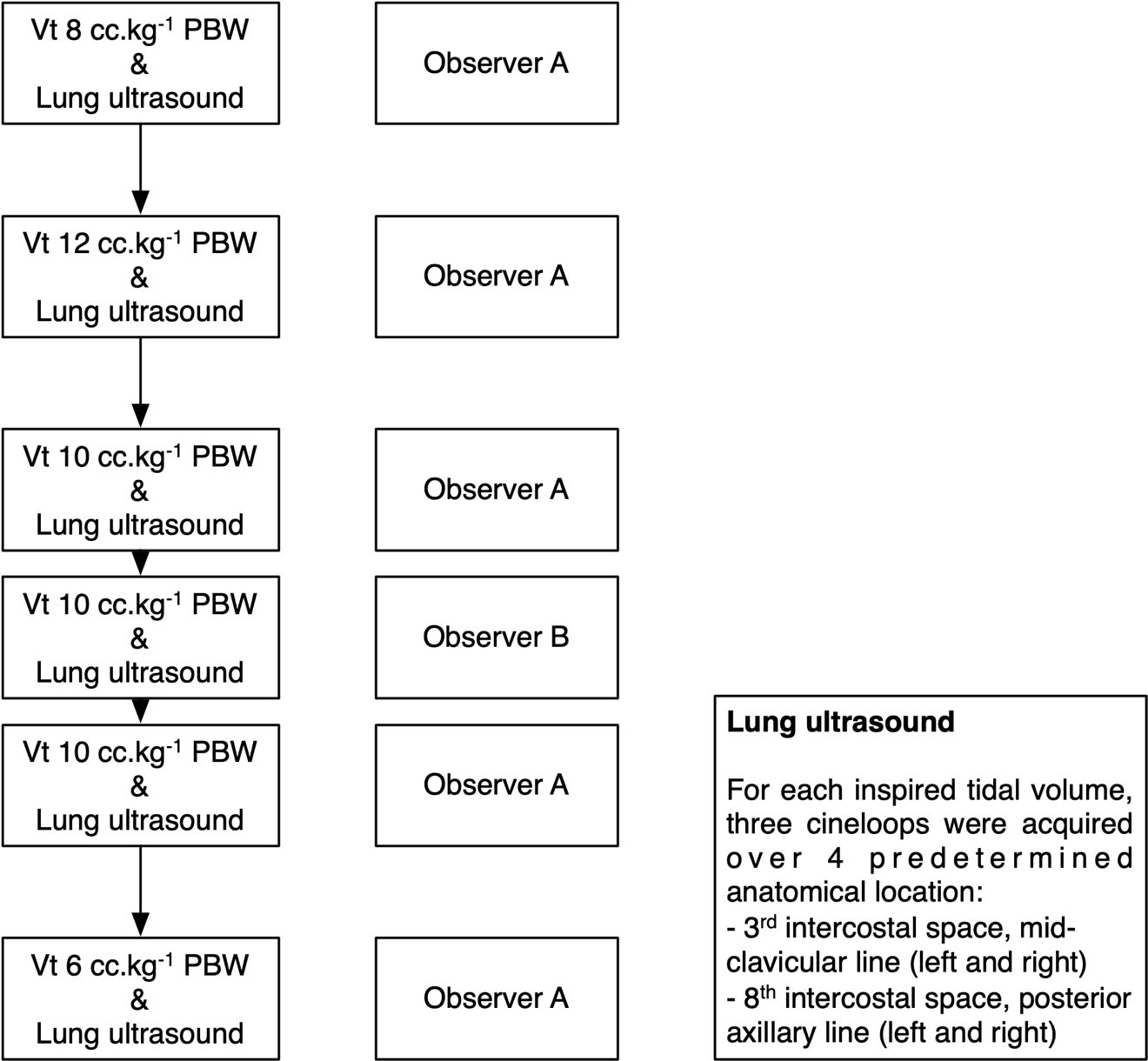

**e-Figure 2**

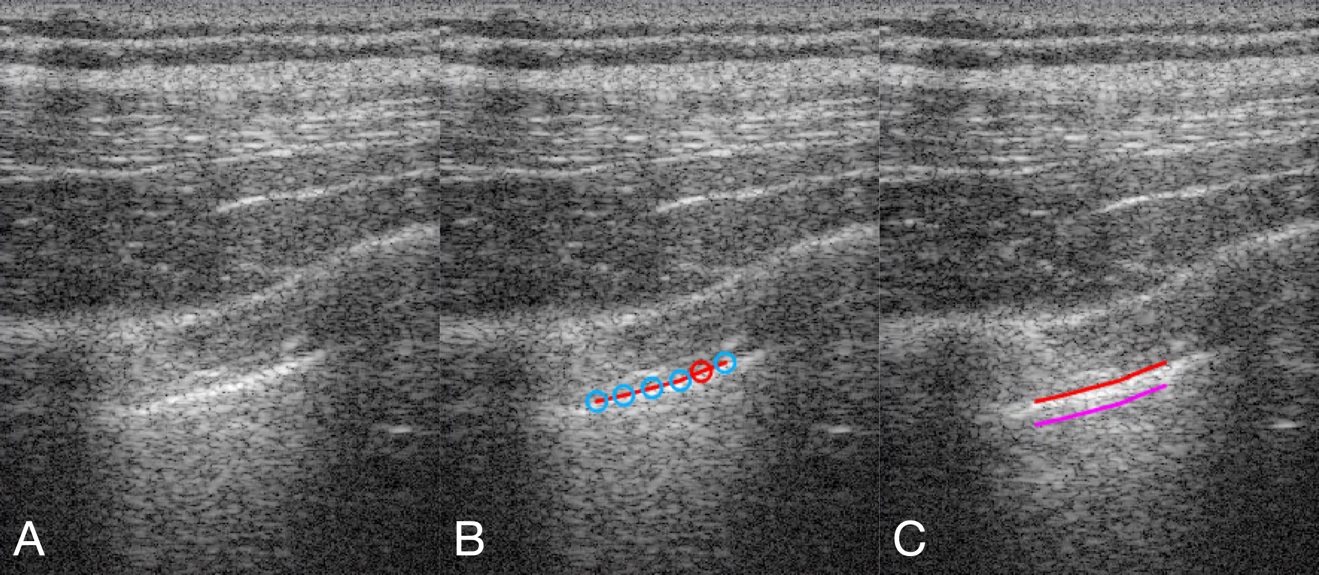

**e-Figure 3**

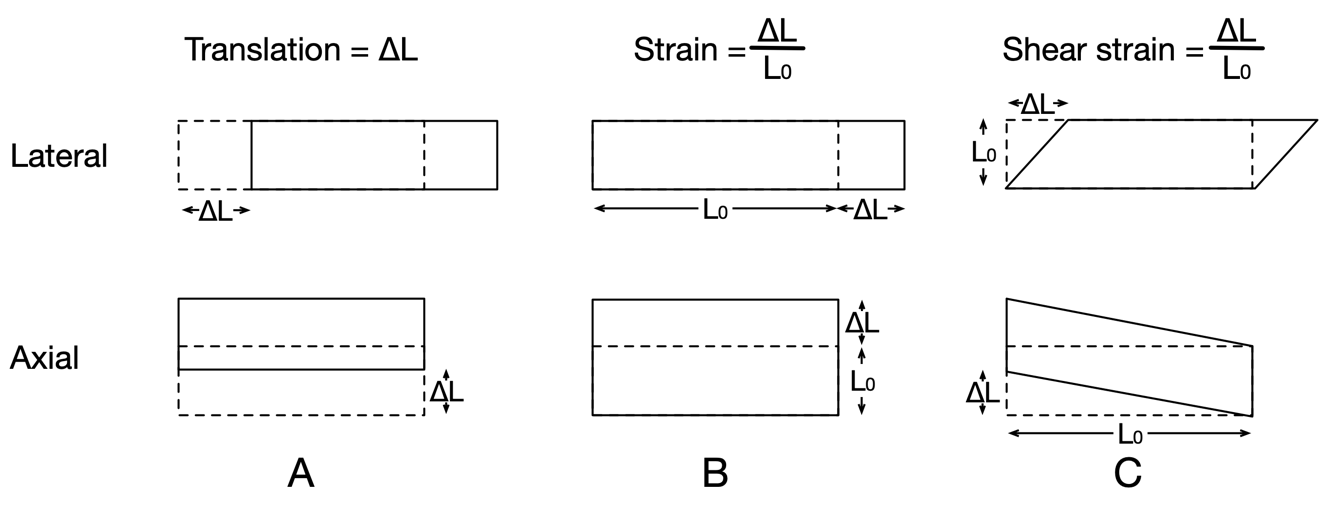

**e-Figure 4**

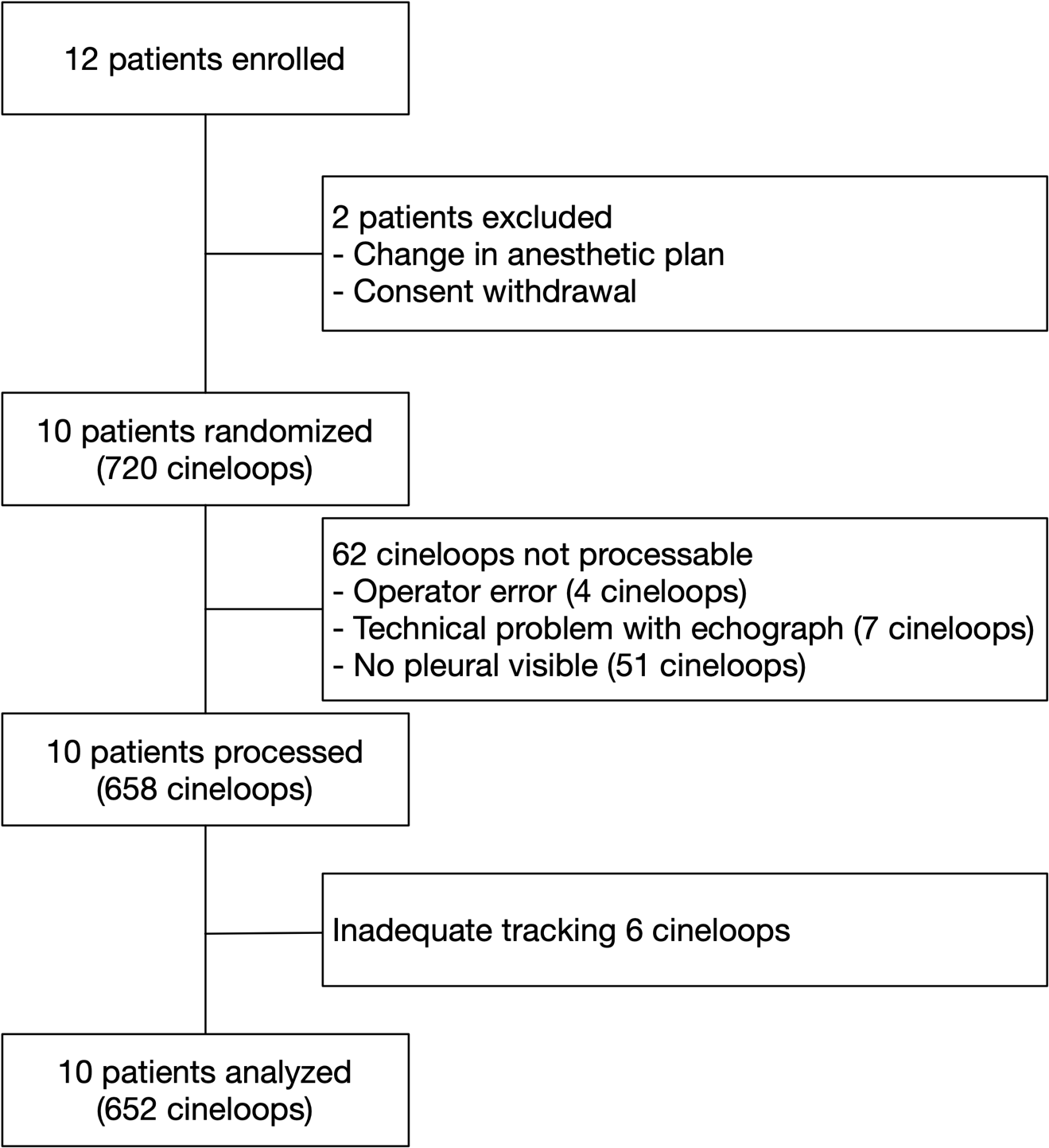

**e-Figure 5**

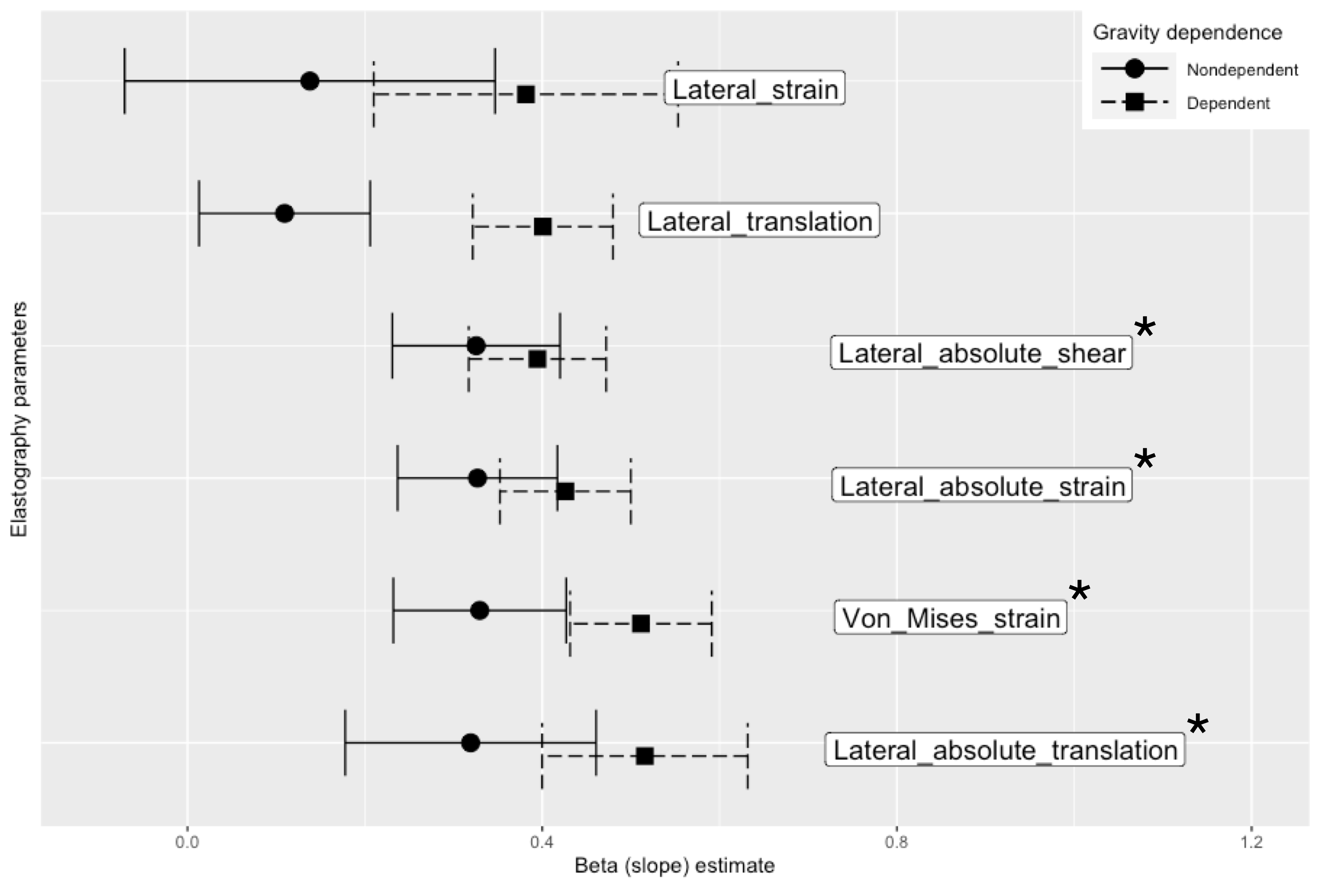

**e-Figure 6**

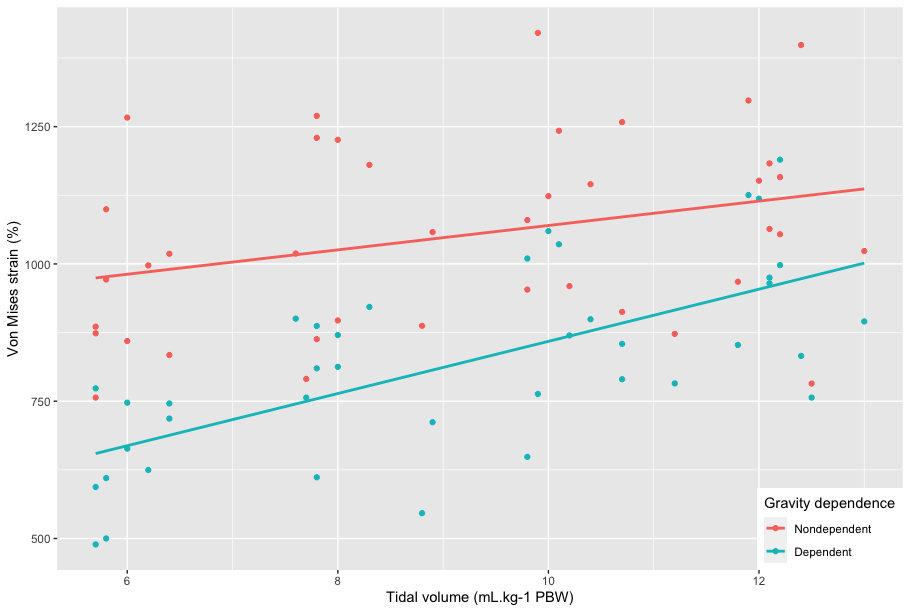

**e-Figure 7**

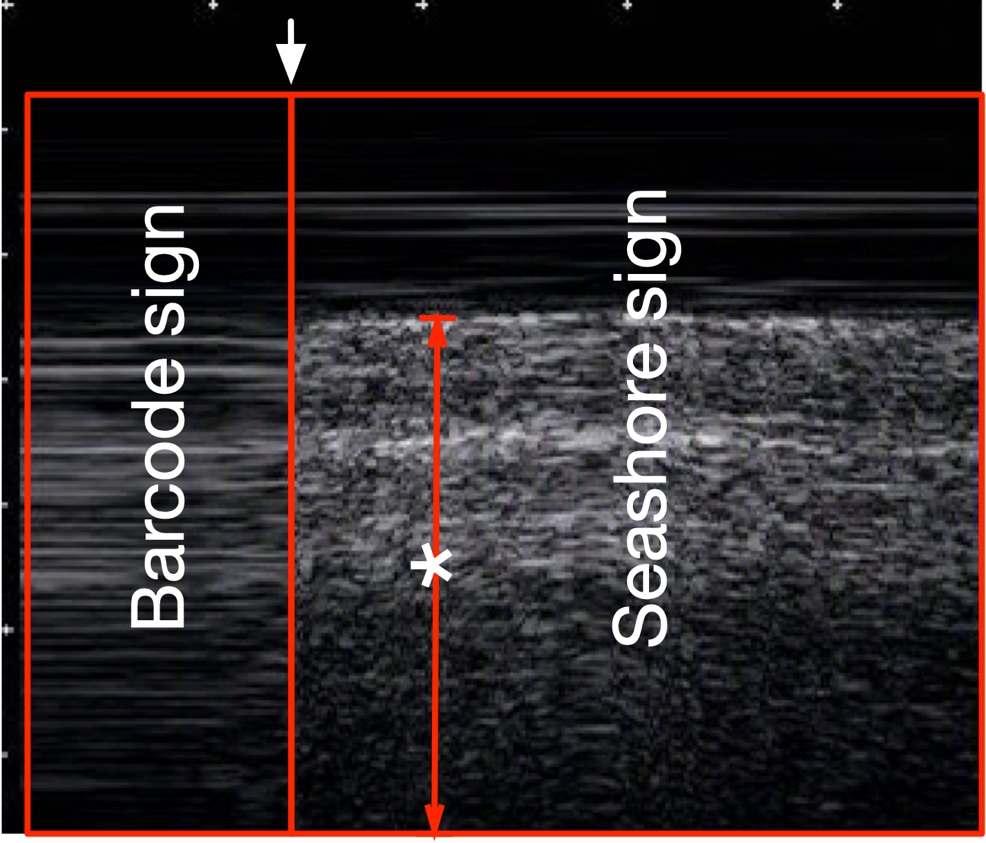

**e-Figure 8**

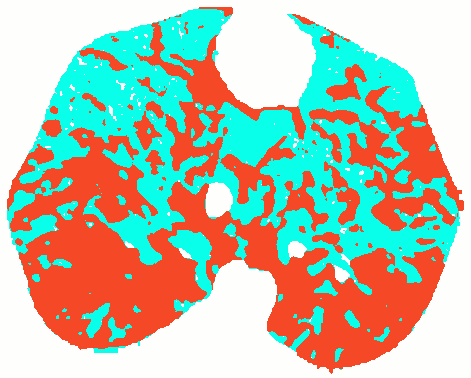
